# Risk Factors for Progression to Alzheimer’s Disease in African Americans in a Large National Cohort

**DOI:** 10.1101/2020.09.17.20196758

**Authors:** Yejin Kim, Sean Savitz, Jessica Lee, Paul E Schulz, Luyao Chen, Xiaoqian Jiang

## Abstract

**Objectives:** To investigate risk factors for progression to Alzheimer’s disease and related dementias (ADRD) in African Americans and non-Hispanic Caucasians in a large US cohort.

**Design:** A matched case-control design using electronic health records (EHRs) from 2000 – 2017.

**Setting:** Cerner EHRs database covering more than 600 Cerner client hospitals. **Participants**: 79,120 patients aged 65 and older (#ADRD=39,560, #non ADRD older adults=39,560) from an initial cohort of 49,826,000 patients.

**Measurements:** We converted ICD9 or ICD10 diagnosis codes into PheWas codes to increase clinical relevance. Then we detected ADRD as having both ADRD diagnosis codes and medications. We considered PheWas codes for Alzheimer’s disease, dementia with cerebral degenerations, senile dementia, and vascular dementia. We considered ADRD medications including acetylcholine and memantine.

**Results:** Using two-step propensity score matching, we built an African American cohort of 4,429 and a 4,570-person matched Caucasian cohort that was similar in terms of onset age, observation length, sex, and known ADRD risks (diabetes, vascular disease, heart disease, head injury, and obesity). Older African Americans had a statistically significant progression from cerebrovascular risk (transient ischemic attack) to ADRD incidence (treatment effect coefficient = 0.0978, *p*-value <0.000) whereas the matched Caucasians did not (treatment effect coefficient = 0.403, *p*-value = 0.196).

**Conclusion:** Our extensive causal analysis using a nationwide EHR discovered disease progression pathways to ADRD. The carefully matched cohorts from different racial groups showed different progression, which partly explains the racial disparities in ADRD incidence.

**IMPACT STATEMENT:** We certify that this work is confirmatory of recent novel clinical research. We confirmed that cerebrovascular disease increases the risk of ADRD incidence more in older African Americans than in non-Hispanic Caucasians when diabetes is controlled.^1,2^ This research adds large-scale and nationwide epidemiological evidence on racial disparities due to cerebrovascular risk.

## INTRODUCTION

Alzheimer’s disease (AD) is the 6th leading cause of death in the United States and it is the only one of the top 10 leading causes of deaths that cannot be cured.^1,2,3^ Currently, at least 5.8 million Americans aged 65 and older suffer from AD.^4^ Alzheimer’s disease and related dementias (ADRD) is a *multifactorial* disease that involves several different etiologic mechanisms with highly heterogeneous phenotypes.^5,6^ Moreover, existing evidence for ADRD suggests that there are significant *disparities* in the prevalence of ADRD across races.^7,8^ Most studies find that older African Americans are more likely than older non-Hispanic whites to be diagnosed with ADRD.^8–11^ Despite some evidence that genetic risk factors for ADRD may differ by race,^12–14^ genetic factors do not appear to account for the large prevalence differences between racial groups.^15,16^ Instead, comorbidity risk factors such as cardiovascular disease, diabetes, and obesity are believed to account for these differences, as they are more prevalent in African Americans.^1,2^

Between racial groups, the influence of comorbid diseases on the incidence of ADRD may differ by their *multifactorial effects*. Thus, it is very important to consider multifactorial influences of comorbid diseases with respect to racial groups to better understand the complicated mechanisms behind health disparities in ADRD. However, such multifactorial analysis must be conducted with extra caution because analyses can easily lead to incorrect conclusions. For example, high levels of education in affluent countries are known to decrease the risk of ADRD, but rising levels of obesity and diabetes may obscure this effect.^17^ Current health disparities studies in ADRD also often focus on hypothesis-testing of single factors for the risk of ADRD, with less attention being paid to multifactorial effects. They have found that the association of sex,^18^ genetic risk factors,^12–14^ SES (e.g., level of education and poverty),^2,19^ and comorbidities (e.g., vascular disease)^1,2^ on ADRD may differ by race.

*Causal inference* is a promising approach that takes the multifactorial effect into consideration in ADRD incidence. Causal inference is one of the most principled approaches to identifying and analyzing multifactorial effects, as it takes other confounders into account to determine causal effects from one risk factor of interest. Recently, causal inference has gained attention in health disparities research in the public health domain.^20,21^ To date, there are few studies that have utilized causal inference in the health disparities of ADRD. One recent study investigated the effects of SES, lifestyle, cardiometabolic and inflammatory factors conditioned on genetic variants and found educational attainment is associated with a reduced risk of ADRD.^22^ As ADRD is a heterogeneous disease with various etiologies, the causal inference requires an extensive set of comorbid diseases to investigate how one disease might lead into ADRD.

Voluminous electronic health records (EHRs) from nationwide hospitals are a rich source for providing comprehensive and longitudinal data on the development of ADRD. Nationwide EHRs also have less bias across health disparity populations, even though the disparity populations are more likely to have bias in general. Causal inference from the nationwide cohorts has a great potential to provide unbiased insights on the yet unidentified ADRD etiology that causes health disparities.

The goal of this study was to investigate a causal structure in nationwide EHRs for disease progression to ADRD in African Americans compared to non-Hispanic Caucasians. Our approach was to build matched cohorts using propensity score matching and to learn causal structure of diseases that progressed to ADRD.

## METHODS

### Database

We utilized Cerner Health Facts, a large clinical database covering EHRs from more than 600 Cerner client hospitals, from 2000 -- 2017, with a total of 49,826,000 inpatients and outpatients.^23^ The data elements included: demographics, encounters, diagnoses, procedures, lab results, medication orders and medication administration. The Cerner Health Facts database contains de-identified electronic health records (EHR) and is subscribed by the University of Texas Health Science Center for research use (https://sbmi.uth.edu/sbmi-data-service/data-set/cerner/). This nationwide EHRs can increase generalizability of our findings.

### Study subjects and variables

As an initial preprocessing, we only included subjects with observation after age of 65, observations longer than 6 months, diagnosis codes, and a timestamp. Then we included a total of 79,120 subjects from amongst ADRD subjects (n=39,560) and non-ADRD subjects (n=39,560) that were approximately 1:1 matched based on ADRD onset age and observation window. That is, for the ADRD subjects, the observation window started from when any diagnosis code was first recorded and ended when first ADRD onset was recorded. For the non-ADRD subjects, we selected subjects that had the closest observation window by matching age at the observation start and finish. Note that we truncated non-ADRD observations after the age when matched ADRD observations ended. This window matching is a crucial step to avoid bias caused by censored observation, such as different ages and observation lengths (e.g., short observation does not mean that one is free of complications).

Variables of interest were all the diseases that were diagnosed within the observation window, which might have potential implications for ADRD onset. We converted ICD9 or ICD10 diagnosis codes into PheWas codes to increase clinical relevance of the billing codes. PheWas ICD code is a hierarchical grouping of ICD codes based on statistical co-occurrence, code frequency, and human review. For more detail, see reference.^24^ We included 1,125 PheWas disease codes that appeared within the observation window in more than 5% of the subjects. In addition, we derived additional risk factor variables for ADRD such as diabetes, vascular disease, heart disease, head injury, and obesity using the PheWas disease codes.^24^ These risk factors were used in the propensity score matching process in later steps.

Outcome of interest was ADRD onset, which we detected as having both ADRD diagnosis code and medication. The ADRD diagnosis codes were PheWas codes for 290.11 Alzheimer’s disease; 290.12 Dementia with cerebral degenerations; 290.13 Senile dementia; and 290.16 Vascular dementia. The ADRD medications were cholinesterase inhibitors (donepezil, galantamine, rivastigmine) or memantine.

### Two comparable cohorts from African Americans and Caucasians

We built two separate cohorts from African Americans and non-Hispanic Caucasians and compared the two cohorts. Traditional approach is to consider race as a risk variable and combine the two racial groups, but this is not desirable in this study due to different subject sizes. As the number of Caucasians are significantly larger than African Americans, the African Americans have less statistical power on the disease progression findings, and consequently our findings are dominated by this largest racial group.

Thus, our approach was to build comparable cohorts from each racial group. Several steps were taken to avoid selection bias in building the study cohort of African Americans and non-Hispanic Caucasian subjects from the large-scale, heterogeneous database. Direct comparison between African Americans and non-Hispanic Caucasians would produce biases due to differences in population size, ADRD incidence rate, onset age, and sex distributions among the different racial groups.^25,26^ Also, different comorbid diseases, such as obesity, diabetes, heart/vascular disease, and head injury in the African Americans and Caucasians, can cause different level of risk for ADRD onset.

We extracted a pair of cohorts in which patient characteristics followed similar distributions (in terms of age, sex, and known risk factors) except race and other disease history, so that we can solely focus on disease progression path without the confounding effects of demographic profiles and known high risk medical conditions. So, we matched one African American patient to one Caucasian patient based on age, sex, ADRD incidence, and known risk factors (heart disease, vascular disease, head injury, diabetes, obesity, smoking, and alcohol-drinking) using propensity score matching (PSM). PSM is used to align patients on the variables of interest based on their estimated propensity score (or probabilities) of eligibility for cohort selection. This matching process had two steps: i) matching cases (ADRD) and controls (non ADRD) within racial groups and ii) matching African Americans and Caucasians (Fig. 1). As a first step, we matched cases (ADRD) and controls (non-ADRD) subjects according to age and observation window within each racial group. As the incidence of ADRD differs by age, a key factor in avoiding the influence of age was to select control patients (non ADRD) that had similar ages during the observation period. This step was similar to the initial ADRD and non-ADRD matching but was done in each racial group. As a next step, we matched the two racial groups pairwise. We matched patients based on sex and known risk factors in addition to the age distribution. This two-step approach can reduce confounding effects within and between racial groups, so that all the four groups have similar age at observation starts and ends, sex, and known high-risk conditions.

**Figure 1.**
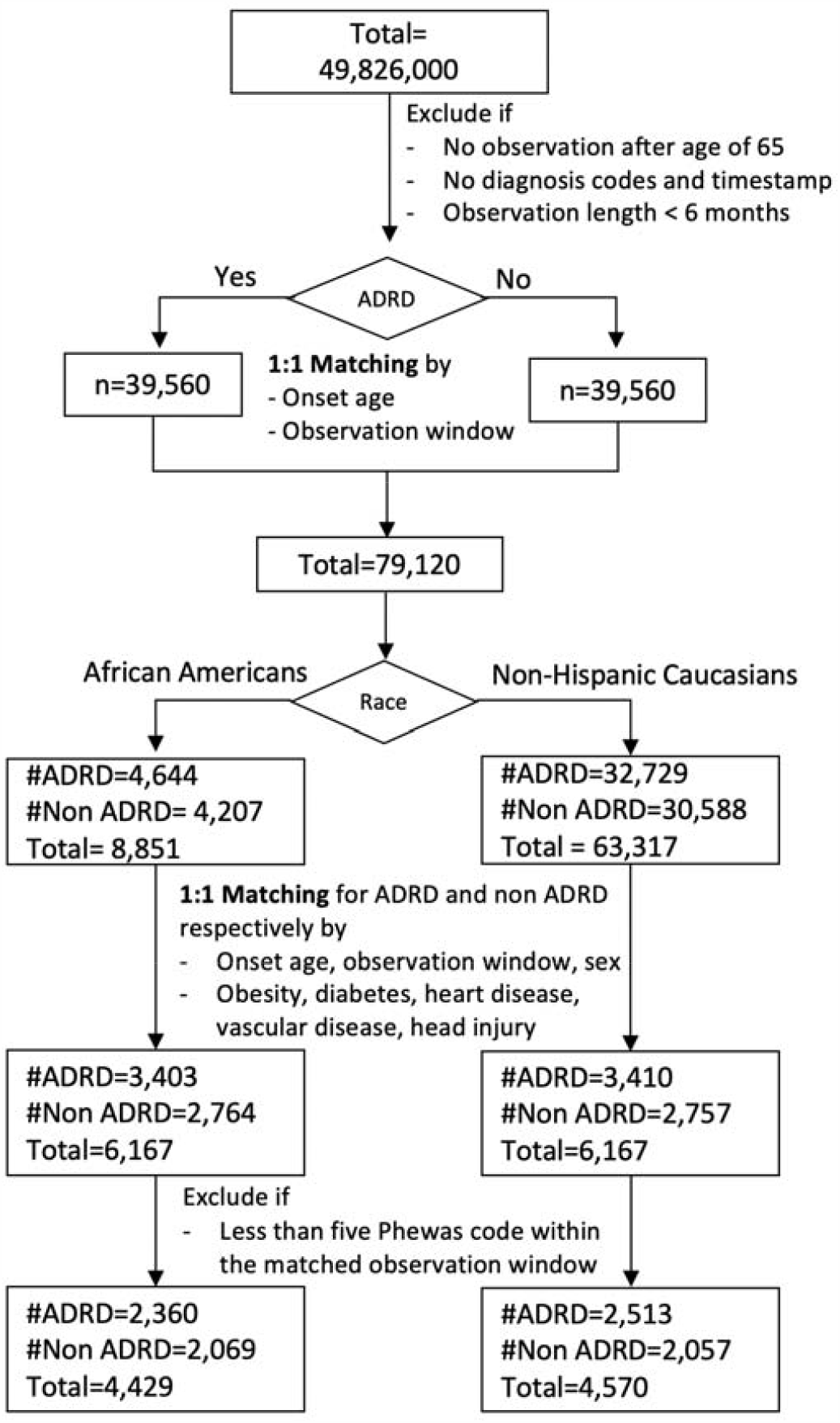
Cohort selection process using propensity score matching to reduce confounding effects.

### Causal structure learning of observed diseases and ADRD

Using the matched cohorts of African Americans and non-Hispanic Caucasians, we investigated and compared how other diseases develop to ADRD for each racial group. We used causal structure learning to find a series of causally related underlying diseases that eventually progressed to ADRD onset, which is called a pathway to ADRD. Causality is used to predict the effects of manipulations.^27^ Understanding cause-effect relationships between variables is of primary interest in many fields. Although intervention in randomized trials is the gold standard to find these relationships, inferring causal information from observational data is a promising alternative approach. Learning Bayesian networks is one way of representing causal structure. It has a form of directed acyclic graph (DAG) whose nodes are random variables and edges refer to conditional dependency. The causal graph naturally considers multifactorial effects by examining conditional dependence or independence given other factors. The causal inference consists of two steps: searching a set of causal graphs (i.e., structure learning) and predicting the effects of a manipulation from the causal models. The causal model search can be very complicated as the number of possible DAGs grows super-exponentially with the number of nodes. A constraint-based search uses conditional independence from data to find *d*-separation, and PC is one of the most popular constraint-based algorithms with moderate accuracy.^28,29^ We applied the PC algorithms to the pairwise cohorts respectively to investigate the differences in causal pathways. We added prior knowledge on temporality during the structure learning: {sex}->{chronic diseases}->{comorbid diseases before ADRD diagnosis}. We encoded the diagnosis histories record as a binary matrix. We used *Tetrad*, a publicly available causality software.^30^ Detailed methods are described in Supplementary Text S1.

### Causal effects

Once we learned the causal structure between diseases before ADRD (unknown risk factors) for each racial group, we derived pathways of the diseases that developed to ADRD. To further validate the causal effect of the pathways, we computed the *treatment effect* to ADRD onset. The treatment effect of treatment, interventions, or exposures in observation data can be assessed as a linear regression model on the causal structure.^31^ To further validate the causal effect, we replaced the risk factors with a random variable to simulate placebo effect. The random variable was a random permutation of values in the risk factor. We repeated 100 times drawing the simulated placebo variable and measured the average coefficient in the linear regression model. We used *Dowhy*, a publicly available package to measure causal effects.^32^

## RESULTS

### Cohort selection

We built matched cohorts of African American and non-Hispanic Caucasian. Of the 49,826,000 patients (Fig. 1) there were 157,620 subjects with ADRD diagnosis codes; 163,320 subjects with ADRD medication codes; and 58,903 subjects with both the diagnosis and medication codes. After excluding subjects without diagnosis/medication codes, timestamp, and observation length less than 6 months, we selected 39,560 ADRD subjects (case) and matched 39,560 non-ADRD subjects (control). We selected African Americans and Caucasians, then performed two-step propensity score matching within and between the racial groups. After the extensive matching and reducing confounding effects, the final cohort was 2,360 ADRD and 2,069 non-ADRD for African Americans; 2,513 ADRD and 2,057 non-ADRD for non-Hispanic Caucasians. This African American cohort and matched Caucasian cohort have a similar number of sample sizes, and follow similar distributions in terms of age, sex, and confounding risk factors (Table 1, Fig. 2).

**Table 1.** Subject’s demographics. Race, age and sex distributions before and after matching. Observation ends at ADRD onset for ADRD patients.

|  | Before |  |  |  | After |  |  |  |
| --- | --- | --- | --- | --- | --- | --- | --- | --- |
|  | African American |  | Caucasian |  | African American |  | Caucasian |  |
|  | ADRD | Non ADRD | ADRD | Non ADRD | ADRD | Non ADRD | ADRD | Non ADRD |
| Total number | 4644 | 4207 | 32729 | 30588 | 3403 | 2764 | 3410 | 2757 |
| Female | 3166 | 2887 | 20400 | 19041 | 2383 | 1953 | 2360 | 1955 |
| Male | 1478 | 1320 | 12329 | 11547 | 1020 | 811 | 1050 | 802 |
| Mean age at observation end (std) | 82.01<br>(6.54) | 84.56<br>(6.76) | 83.45<br>(6.03) | 82.87<br>(5.95) | 82.80<br>(6.21) | 85.06<br>(6.32) | 83.17<br>(6.02) | 84.90<br>(6.18) |
| Diabetes | 1337 | 1127 | 6639 | 7774 | 611 | 509 | 519 | 488 |
| Heart disease | 1110 | 1073 | 8804 | 11713 | 561 | 597 | 493 | 595 |
| Vascular disease | 1075 | 974 | 8269 | 10977 | 549 | 551 | 483 | 558 |
| Obesity | 168 | 307 | 956 | 1931 | 41 | 77 | 24 | 57 |
| Head injury | 126 | 183 | 1619 | 1350 | 43 | 58 | 39 | 33 |

**Figure 2.**
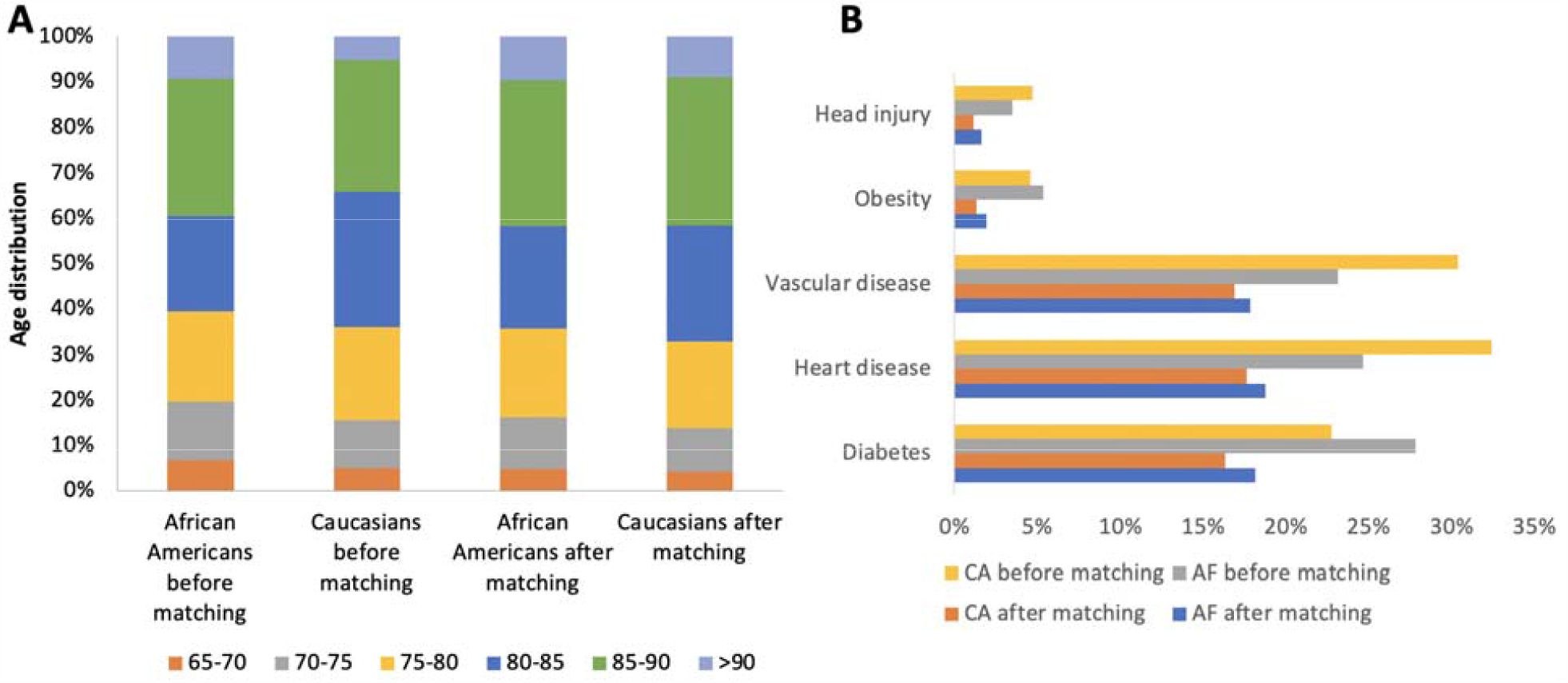
Subject’s age and known risk factor distributions. (a) Onset age distribution of ADRD patients after matching Caucasians to African Americans. Onset age of (original) Caucasians tends to be older than that of African Americans. The matched Caucasian follows a similar distribution. (b) Distribution of known risk factors after matching. The matched Caucasian follows a similar distribution with African American in terms of the confounding risk factors. Ca = Caucasian, Paired Ca = Caucasian that are matched to African American. AF= African American.

### Causal structure of observed diseases and ADRD

We derived a causal structure of commonly observed 1,125 diagnoses for each racial group. Note that we analyzed the pathways with minimized effects of known risk factors (diabetes, vascular disease, heart disease, head injury, obesity), so that they are discouraged to appear in the pathways. In the causal structure learning, we learned two separate causal graphs for African American and its matched Caucasian. As a result, we found that the two causal structures from African American and its matched Caucasian shares similar but also distinctive pathways, even though the two cohorts follow similar distributions in age, sex, and other risk factors (Fig. 3). Both racial groups shared similar psychological/cognitive disorders pathways (note that we omitted precedent variables due to limited space):

**Figure 3.**
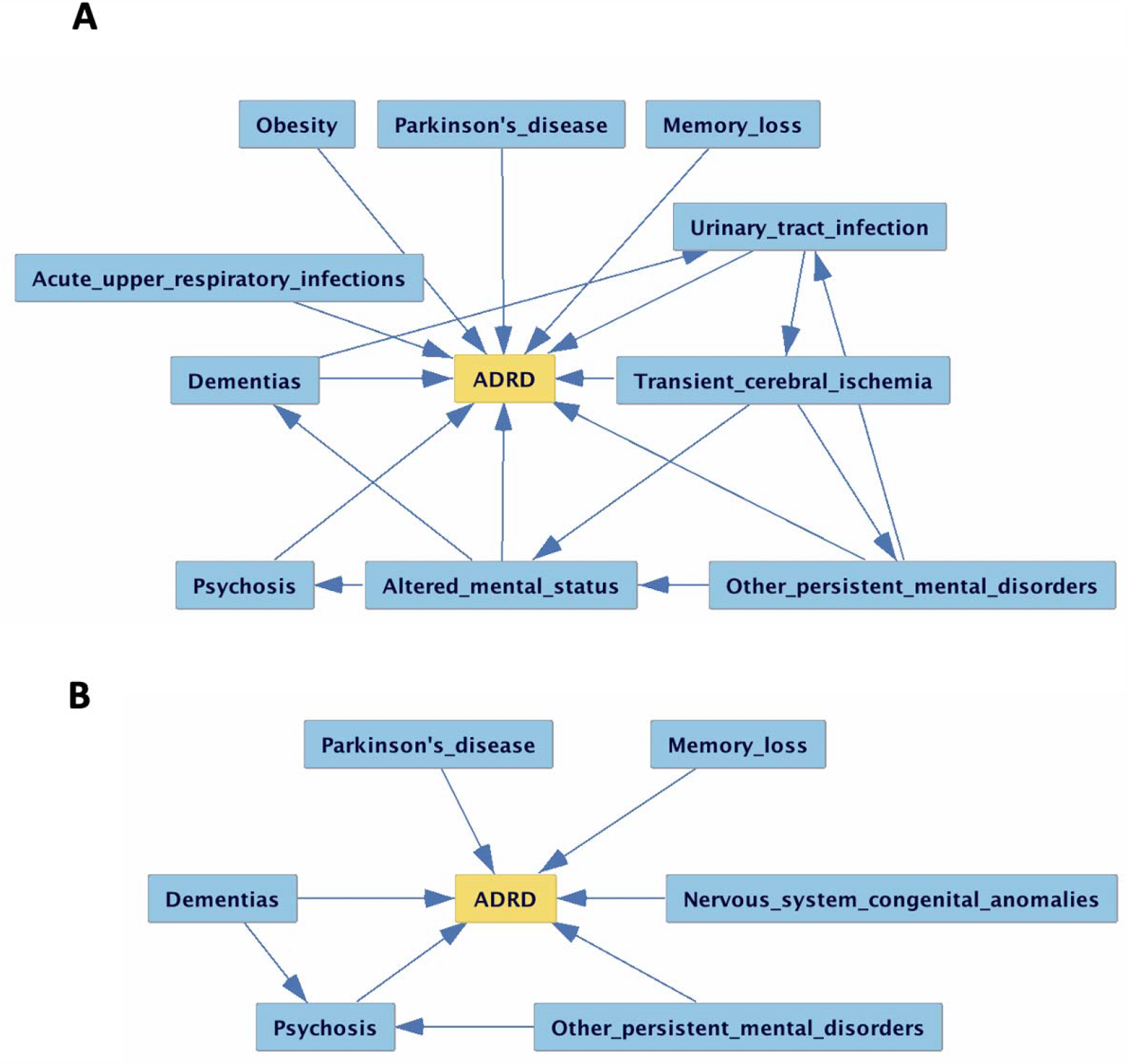
Disease progression pathways leading to ADRD onset in (a) African Americans and (b) matched Caucasians. Due to limited space, we only presented Markov blankets for ADRD. Compared to Caucasians, African Americans had a stronger cerebrovascular pathway: {Type2 diabetes/chronic vascular disease -> *Transient cerebral ischemia -> ADRD*} and {*Aphasia* <-*Transient cerebral ischemia -> ADRD*}

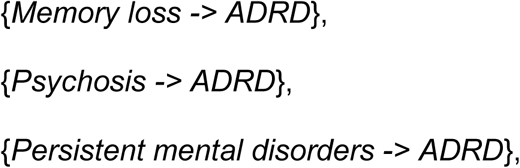

and other neurodegenerative pathways:

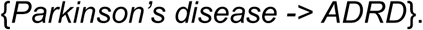

African Americans had a distinctive cerebrovascular disease pathway:

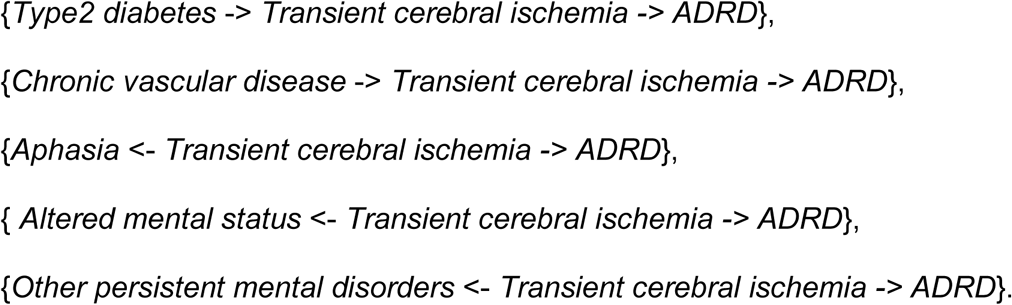

African Americans also had distinctive pathways:

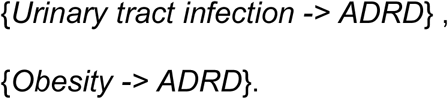

We measured the causal effect of transient cerebral ischemia on the incidence of ADRD (Table 2). As a result, transient cerebral ischemia had a statistically significant positive causal effect on ADRD incidence in African Americans (coefficient = 0.0978, *p*-value <0.000 in Table 2); whereas there was no significance in Caucasians (coefficient = 0.403, *p*-value = 0.196). Causal effect of the placebo variables (in regard to the transient cerebral ischemia) was coefficient=0.0007 and *p*-value = 0.48 (Table 2).

**Table 2.**
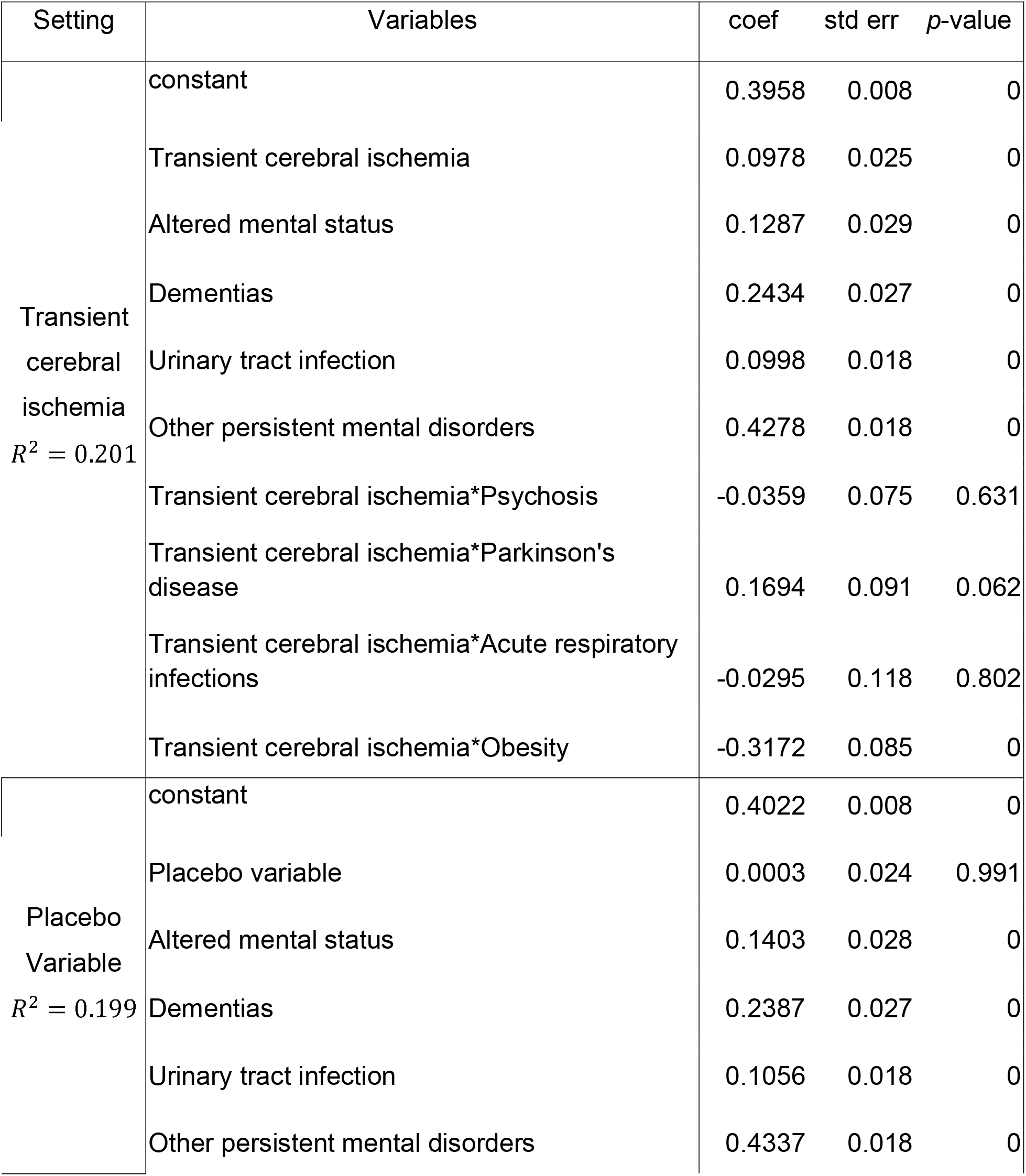
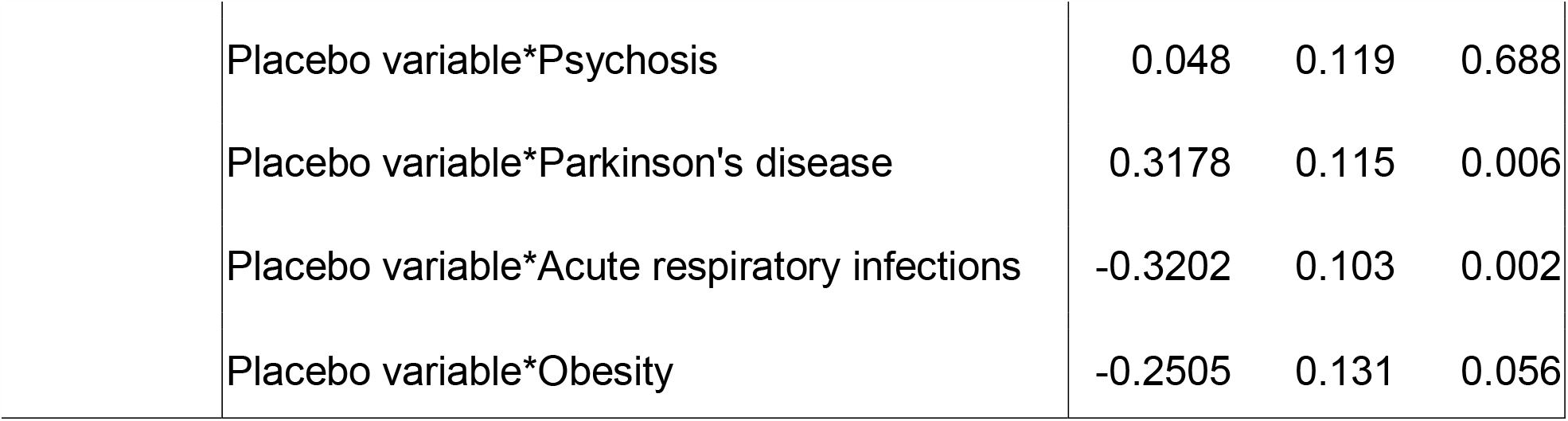
Causal effect of transient cerebral ischemia and placebo variable to ADRD onset measured by linear regression coefficient. We replaced transient cerebral ischemia with a placebo variable generated by random permutation of transient cerebral ischemia.

## DISCUSSION AND CONCLUSIONS

The objective of this study was to investigate different disease progressions to ADRD in African Americans compared to non-Hispanic Caucasians using diagnosis codes in a nationwide EHR. Using two-step propensity score matching, we selected a matched Caucasian cohort that had a similar distribution with an African American cohort in terms of ADRD onset age, age at observation begins and ends, sex, prevalence of diabetes, vascular disease, heart disease, head injury, and obesity. Using PC causal structure learning, we derived a series of diagnoses that eventually lead to ADRD. Then we tested the statistical significance of the pathway’s causal effect on ADRD onset. We believe that our causal inference to derive a disease progression pathways was rigorous in that we i) included both people with and without ADRD in the same age to reduce bias, ii) assessed a wide range of potential risk factors apart from known ones,^33^ and iii) maximized the clinical relevance of billing codes by converting the ICD9 or ICD10 codes to PheWas codes.^24^

In our study, we found that there are several significant differences between the disease progression pathways to ADRD in African Americans when compared to Caucasians. African Americans had statistically significant cerebrovascular pathways than the matched Caucasians, which implies that the cerebrovascular diseases of African Americans were more likely to progress to ADRD than that of Caucasians.

Our finding on the progression of cerebrovascular diseases to ADRD in African Americans may explain why ADRD is reported more frequently in Africans Americans.^34–38^ In addition to the fact that transient cerebral ischemic attack (TIA) is more prevalent in African Americans,^39^ we found that TIA is more likely to progress to ADRD in African Americans than Caucasians. TIA may be a sign of strokes causing vascular dementia,^40,41^ and consequently contributes to the higher incidence of ADRD in African Americans. Please note that chronic vascular disease and diabetes, consistent causes of TIA, were controlled in ADRD and non-ADRD subjects in both racial groups (Table 1), but still we observed TIAs more progress to ADRD in African Americans. These findings suggest that TIAs may be a controllable risk factor that can reduce the risk of ADRD onset in African Americans.

Taken together, these findings suggest that extensive causal analysis from nationwide EHRs can demonstrate the disease progression pathways to ADRD. Moreover, comparing the carefully matched cohorts from different racial groups allowed us to examine possible explanations of racial disparities in ADRD incidence. In particular, we observed that TIAs may be a controllable risk factor for ADRD onset in African Americans.

This study inevitably inherits the limitation of data quality in EHRs. In spite of an advantage of the EHRs that they can extensively cover a population’s long medical history in breadth, EHRs are mainly collected for billing, not for studying, and thus diagnosis billing codes in EHRs are sometimes incomplete and lack detail in depth. EHRs cannot capture many socioeconomic status (e.g., education, literacy, life course exposures), which are one of the main causes of racial disparities in ADRD.^16,42^ Also the population in the Cerner EHRs is mostly from the middle class that is privately insured, which might not fully represent the general population in the US. Another limitation is that ADRD is a progressive disease that shows slow development over the several years to decades. EHRs might not be able to fully capture the long progression. Late ADRD onset age (due to social determinants) also showed the limitation of EHRs. Also the PSM method can potentially cause bias as the unmatched patients were excluded in the analysis. Notwithstanding these limitations, this study offers some important insights into multifactorial mechanisms of racial disparities in ADRD onset.

## Data Availability

We used deidentified Cerner EHRs that University of Texas Health Science Center at Houston subscribes.

## Conflict of Interest

There is no conflict of interest

## Author Contributions

Y.K., J.L., P.S., and X.J. participated in writing the manuscript;

Y.K., L.C., and X.J. participated in the experiment design and implemented the statistical analysis;

J.L. and P.S. provided clinical perspectives on the findings.

## Sponsor’s Role

YK was supported in part by UTHealth startup, UT Stars award, and the National Institute of Health (NIH) under award number R01GM124111. XJ is CPRIT Scholar in Cancer Research (RR180012), and he was supported in part by Christopher Sarofim Family Professorship, UT Stars award, UTHealth startup, and the National Institute of Health (NIH) under award number R01GM114612 and U01TR002062. PES is supported by NIH, The Weston Brain Institute, The Kleberg Foundation, donations, and numerous pharmaceutical companies.

All sponsors did not play a role in the design, methods, subject recruitments, data collections, analysis, and preparation of paper.

## Notes

### Competing Interest Statement

The authors have declared no competing interest.

### Author Declarations

This work is reviewed by IRB at University of Texas Health Science Center at Houston (HSC-SBMI-13-0549)

